# Perceived facilitators and barriers to physical activity, using the COM-B Model for Behavioural Change, in people with chronic pain: a qualitative evaluation of patient and stakeholder perspectives

**DOI:** 10.1101/2024.12.11.24318767

**Authors:** C Higgins, PM Dall, C Leese, P Adair, PA Cameron, SK Inglis, A Christogianni, BH Smith, L Colvin

## Abstract

**Background:** Physical inactivity is a substantial public health concern and a particularly challenging problem to address. Physical activity is shown to be of therapeutic benefit to those living with chronic pain, but intervention studies have often been unsuccessful in promoting sustainable engagement with physical activity in this population.

**Aim:** To develop a more comprehensive understanding of the barriers to engaging with sustainable physical activity and the facilitators that promote this engagement in people with chronic pain.

**Design & setting:** The study design was qualitative, informed by behaviour change theory (the COM-B model), and participants were drawn from two settings. First, 36 adults living with moderate-severe chronic pain were recruited from a specialist NHS pain service. Secondly, 15 stakeholders were recruited to represent a range of healthcare sectors involved in supporting people with chronic pain.

**Method:** One-to-one semi-structured interviews, informed by the COM-B model, were conducted between May 2020 and October 2021. Interviews were conducted by telephone or using online conferencing software and were transcribed *verbatim* and anonymised. The Framework Method was used to interpret the data.

**Results:** The barriers to sustainable physical activity engagement in people with chronic pain are expansive, and the inclusion of stakeholder perspectives is valuable in developing a more comprehensive understanding of these barriers and also of the facilitators that are successful in promoting sustainable physical activity in this population.

**Conclusions:** The COM-B model provides a robust theoretical framework for organising and understanding these barriers and facilitators in a way could that inform effective intervention design.

## Introduction

For more than a decade, physical inactivity has been widely recognised as one of the greatest public health concerns of the 21^st^ century (Blair 2009; Kohl *et al*., 2012; Rütten *et al*., 2013; Trost *et al*., 2014), and studies show that this is a particular issue in people living with chronic pain (CP) (Fjeld *et al*., 2023; McBeth *et al*., 2010; Parker *et al*., 2017). Physical inactivity is generally considered to be activity levels that falls below those recommended in current guidelines. Advice from the World Health Organization (2020) and the UK’s Chief Medical Officers (2019) recommends at least 150 minutes per week of moderate activity or at least 75 minutes of vigorous activity. In people with CP, physical inactivity is shown to increase risk of mortality (Smith *et al*., 2018), while physical activity (PA) is shown to reduce pain severity/sensitivity (Geneen *et al*., 2017; Rice *et al*., 2019), increase pain tolerance (Årnes *et al*., 2023) and improve quality of life (Amiri, 2022; Geneen *et al*., 2017; NICE, 2021). Many PA interventions have been trialled; however, the evidence for their success in people with CP is generally weak. Indeed, several recent systematic reviews have shown no effect of interventions at increasing sustainable PA in this population (Booth *et al*., 2022; Oliveira *et al*., 2016; Marley *et al*., 2017). In consequence, there is an urgent need to develop a comprehensive understanding of the barriers and facilitators to PA in people with CP, particularly within the context of a behavioural change model to better inform effective interventions.

Over the past two decades, in response to a growing recognition of the importance of PA in the management of chronic, painful conditions, qualitative studies have attempted to develop an understanding of the barriers to engaging with PA and the facilitators that might support such engagement (Boutevillain *et al*., 2017; Dnes *et al*., 2020; Joelsson *et al*., 2017; Kanavaki *et al*., 2017; Meade *et al*., 2021; Vader *et al*., 2021). Most qualitative studies have focused solely on patient perceptions; however, there is demonstrated value in also including stakeholder perspectives (Lindner *et al*., 2023) and in interpreting the data within a model of behavioural change, such as the COM-B model, to maximise the potential of developing effective interventions (Webb *et al*., 2022).

The COM-B Model for Behavioural Change (Michie *et al*., 2011) is centred on a system of behaviour and is extended to capture the range of mechanisms that could be involved in change, both internal (psychological and physical factors) and external (environmental factors). At the core of the model (the behaviour system), they propose that an individual must have the capability (C), opportunity (O) and motivation (M) in order to achieve a behaviour (B) and that one or more of these components must change in order to achieve behavioural change. They acknowledge that these factors interact over time; therefore, behaviour is considered a dynamic system with positive and negative feedback loops. The authors provide a visual representation of their model, referred to as the ‘behaviour change wheel’ (BCW; see Figure 1; Michie *et al*., 2011).

**Figure 1:**
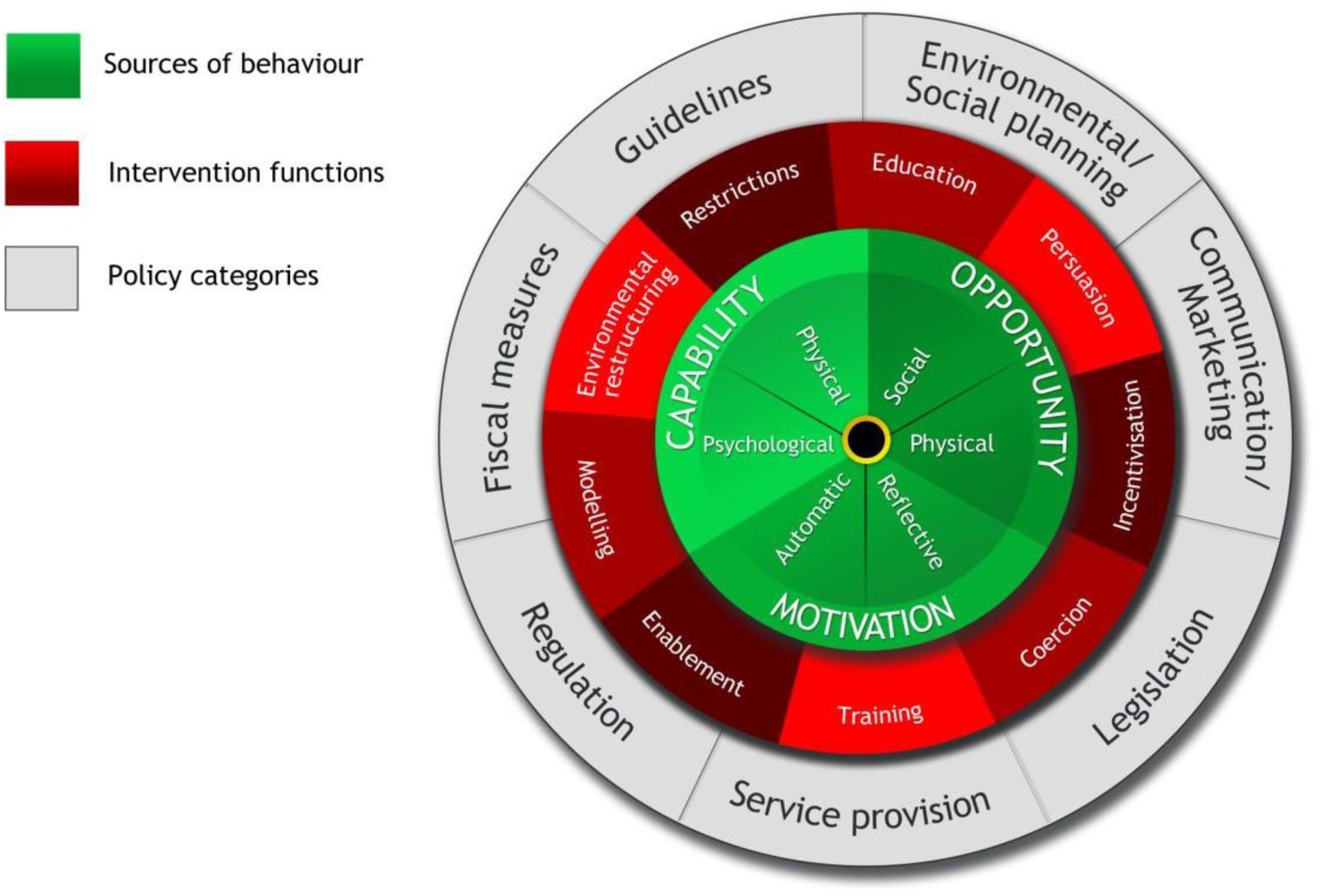
Behaviour Change Wheel (BCW) from the COM-B Model for Behavioural Change [Reproduced from Michie *et al*. (2011), with permission from Springer Nature (2 January 2024).]

The present study set out to understand the barriers and facilitators to increasing PA for those living with CP with the aim of gathering evidence to produce a SUstainable Self Effective Exercise Development (SUSSED) intervention to support clinical decision-making.

## Methods

The study design was qualitative, informed by behaviour change theory (the COM-B model), and participants were drawn from two settings. First, 36 adults living with CP (hereafter termed ‘patients’) were recruited from a specialist National Health Service (NHS) pain service in Scotland. NHS healthcare is available to all in the country and is free at the point of contact, and access to specialist pain services requires a referral from a general practitioner (GP), secondary care clinician or other appropriately qualified healthcare professional. A specialist setting was selected to ensure the recruitment of patients with a range of severe and debilitating painful conditions, such as fibromyalgia, osteoarthritis and chronic back pain. Patient inclusion criteria were: (1) at least 18 years of age; (2) consented to being contacted for research purposes and able to give informed consent for study participation; (3) referred to the Pain Service; (4) pain that had persisted for at least 6 months; (5) moderate-severe pain, assessed prior to study inception using a 0-10 numeric rating scale (employing the commonly-used cut-point of >3 (Boonstra *et al*., 2016)); and (6) no clinical contraindications to wearing an accelerometer. Patient participants were excluded if they were in active treatment for cancer or were participating in the clinical phase of an interventional study or had done so within the past 30 days. Pain score (obtained from the numeric rating scale) and PA level (number of steps per day, obtained from ActivPAL accelerometers, which were worn by all participants) are described for each patient participant.

Secondly, 15 stakeholders were recruited nationally to represent a range of healthcare sectors, including GPs, physiotherapists and other service providers involved in supporting patients with CP. Stakeholder inclusion criteria were: (1) at least 18 years of age; (2) able to give informed consent for participation; and (3) considered to be fulfilling a relevant stakeholder role (i.e. GPs and allied health professionals (AHP)s, nurses, pharmacists, carers, staff at third sector organisations, leisure centre staff and others identified through the study team working in conjunction with the Green Health Partnership, Pain Concern and Versus Arthritis). Stakeholders were recruited to provide an alternative insight into the barriers and facilitators encountered by patients living with CP, which was triangulated with the data provided by patients.

One-to-one semi-structured interviews were conducted with participants between May 2020 and October 2021, during the height of the COVID-19 pandemic, and written consent was taken from all participants prior to participation. Interviews were conducted using online conferencing software (Microsoft Teams) or by telephone, due to imposed lockdown restrictions, and the interview topic guides were based on the COM-B model (Michie *et al*., 2011). Interviews were recorded using either an Olympus audio recorder (telephone interviews) or using video conference software (Microsoft Teams), transcribed verbatim and anonymised prior to data analysis.

Using an inductive-deductive approach, the Framework Method (Gale *et al*., 2013) was employed in interpreting the data produced in the present study, and data analysis was conducted using NVivo 12 Plus (Lumivero, 2018). First, inductive line-by-line coding was performed independently by two researchers using three patient transcripts (CH & PA) and three stakeholder transcripts (CH & AC). Secondly, using a consensus approach, these codes were collapsed to identify a preliminary list of subthemes – the initial iteration of the thematic framework. The thematic framework was considered a ‘living document’, which was not finalised until the final transcript had been analysed and all possible subthemes had been identified. Thirdly, the inductively identified subthemes were classified within the deductively identified themes (i.e. the six components of the COM-B model). This final step in the process was conducted using a consensus approach involving three researchers (CH, CL & BHS), with opportunities for input from all team members.

Further information about this study can be found on ISRCTN (ISRCTN95480359, https://doi.org/10.1186/ISRCTN95480359).

## Results

A total of 36 patients and 15 stakeholders participated in interviews, and their characteristics are shown in Table 1. The patient cohort comprised more females (61%; n=22) than males (39%; n=14), and ages ranged from 21 to 81 years (mean = 52±16 years). The mean pain intensity score was 5.03±1.14 on a 0-10 visual analogue scale: 3 (9%) participants reported experiencing ‘mild’ pain (scoring ≤3); 27 (77%) reported ‘moderate’ pain (scoring 4-6); and 5 (14%) reported ‘severe’ pain (scoring ≥7). Number of steps per day, based on accelerometer data, ranged from 702 to 23,945 (mean = 6,881±5,106).

**Table 1:**
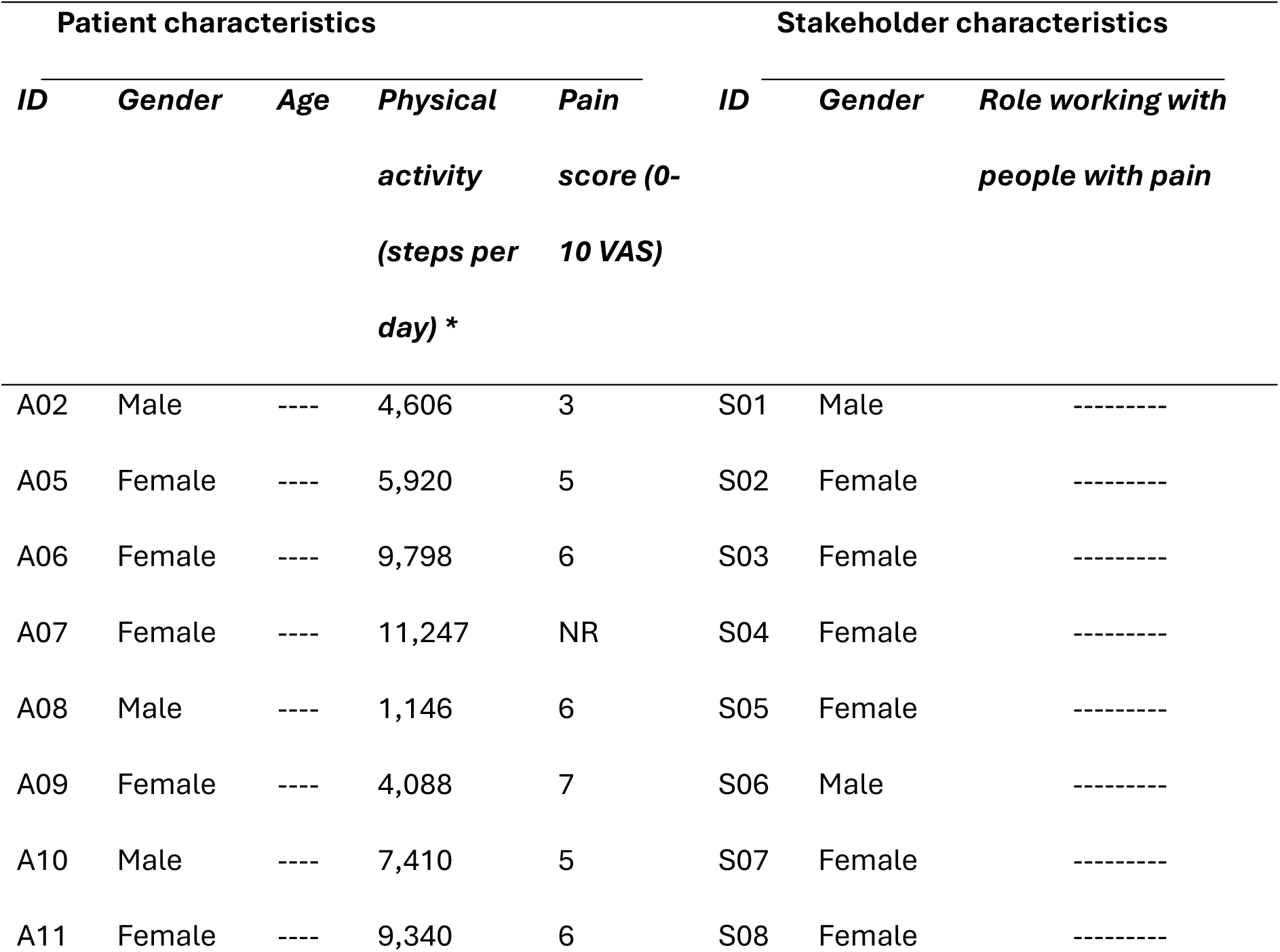
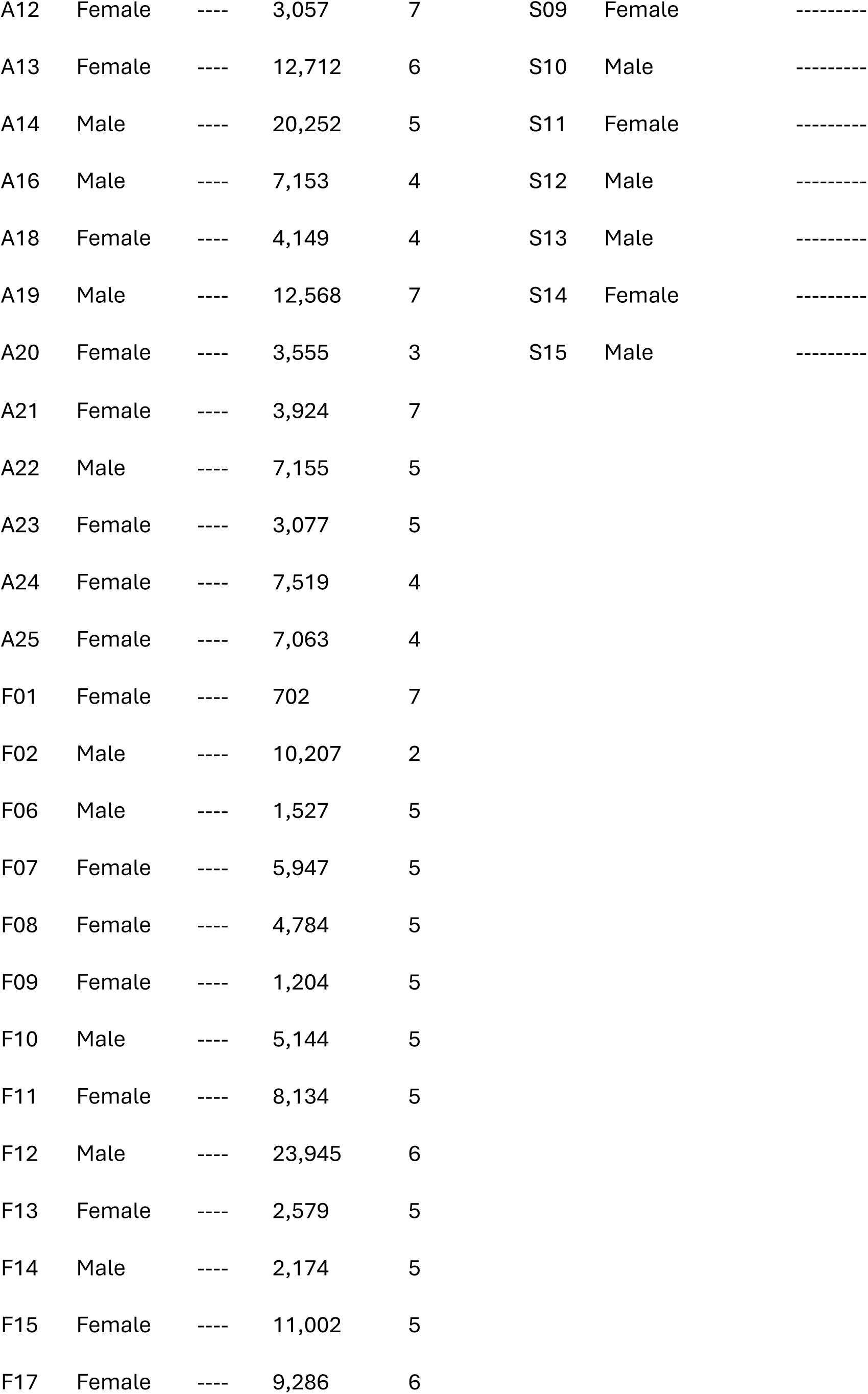
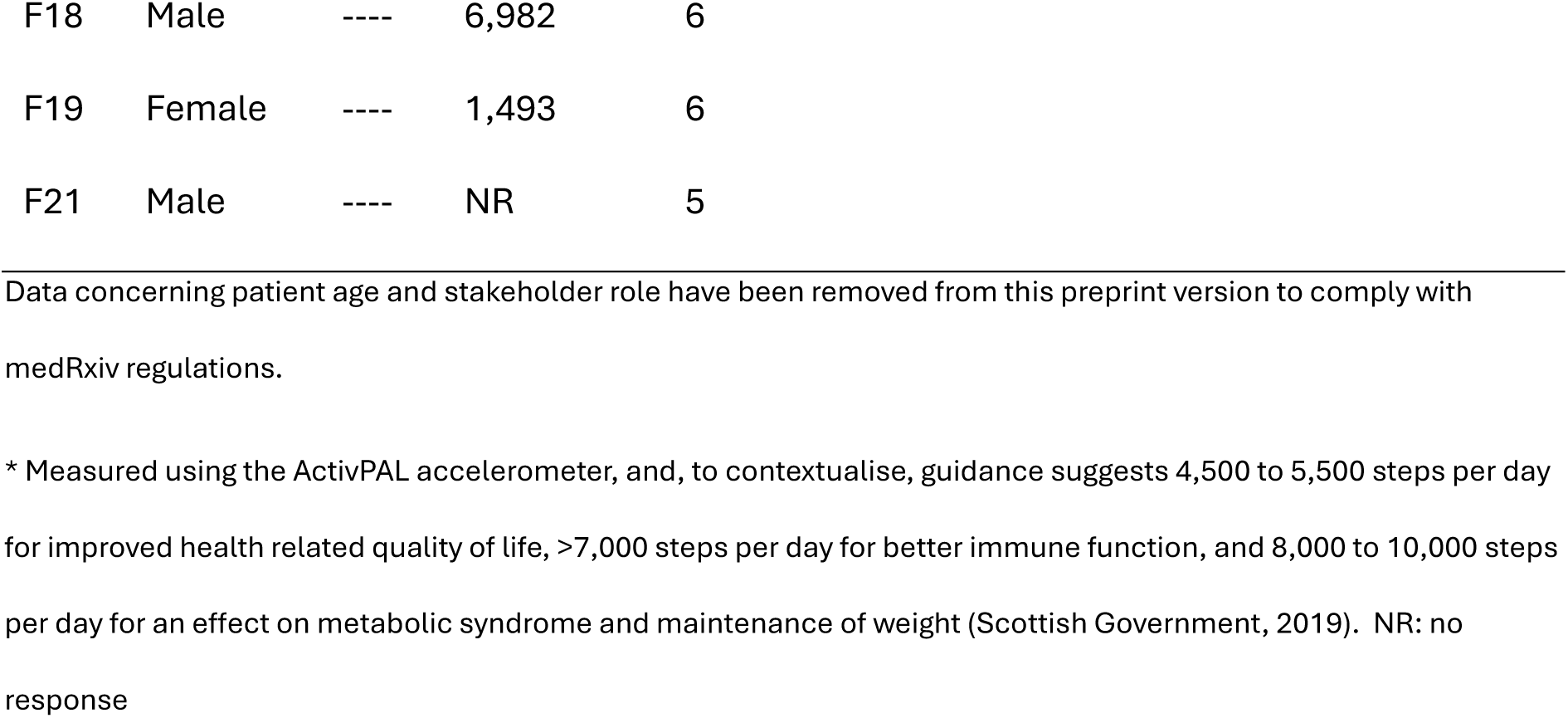
Patient and stakeholder demographic characteristics and measures of pain and physical activity in patient participants.

| Patient characteristics |  |  |  |  | Stakeholder characteristics |  |  |
| --- | --- | --- | --- | --- | --- | --- | --- |
| *ID* | *Gender* | *Age* | *Physical activity (steps per day) \** | *Pain score (0-10 VAS)* | *ID* | *Gender* | *Role working with people with pain* |
| A02 | Male | ---- | 4,606 | 3 | S01 | Male | ----- |
| A05 | Female | ---- | 5,920 | 5 | S02 | Female | ----- |
| A06 | Female | ---- | 9,798 | 6 | S03 | Female | ----- |
| A07 | Female | ---- | 11,247 | NR | S04 | Female | ----- |
| A08 | Male | ---- | 1,146 | 6 | S05 | Female | ----- |
| A09 | Female | ---- | 4,088 | 7 | S06 | Male | ----- |
| A10 | Male | ---- | 7,410 | 5 | S07 | Female | ----- |
| A11 | Female | ---- | 9,340 | 6 | S08 | Female | ----- |
| A12 | Female | ---- | 3,057 | 7 | S09 | Female | ----- |
| A13 | Female | ---- | 12,712 | 6 | S10 | Male | ----- |
| A14 | Male | ---- | 20,252 | 5 | S11 | Female | ----- |
| A16 | Male | ---- | 7,153 | 4 | S12 | Male | ----- |
| A18 | Female | ---- | 4,149 | 4 | S13 | Male | ----- |
| A19 | Male | ---- | 12,568 | 7 | S14 | Female | ----- |
| A20 | Female | ---- | 3,555 | 3 | S15 | Male | ----- |
| A21 | Female | ---- | 3,924 | 7 |  |  |  |
| A22 | Male | ---- | 7,155 | 5 |  |  |  |
| A23 | Female | ---- | 3,077 | 5 |  |  |  |
| A24 | Female | ---- | 7,519 | 4 |  |  |  |
| A25 | Female | ---- | 7,063 | 4 |  |  |  |
| F01 | Female | ---- | 702 | 7 |  |  |  |
| F02 | Male | ---- | 10,207 | 2 |  |  |  |
| F06 | Male | ---- | 1,527 | 5 |  |  |  |
| F07 | Female | ---- | 5,947 | 5 |  |  |  |
| F08 | Female | ---- | 4,784 | 5 |  |  |  |
| F09 | Female | ---- | 1,204 | 5 |  |  |  |
| F10 | Male | ---- | 5,144 | 5 |  |  |  |
| F11 | Female | ---- | 8,134 | 5 |  |  |  |
| F12 | Male | ---- | 23,945 | 6 |  |  |  |
| F13 | Female | ---- | 2,579 | 5 |  |  |  |
| F14 | Male | ---- | 2,174 | 5 |  |  |  |
| F15 | Female | ---- | 11,002 | 5 |  |  |  |
| F17 | Female | ---- | 9,286 | 6 |  |  |  |

|  |  |  |  |  |
| --- | --- | --- | --- | --- |
| F18 | Male | ---- | 6,982 | 6 |
| F19 | Female | ---- | 1,493 | 6 |
| F21 | Male | ---- | NR | 5 |

Data concerning patient age and stakeholder role have been removed from this preprint version to comply with medRxiv regulations.
\* Measured using the ActivPAL accelerometer, and, to contextualise, guidance suggests 4,500 to 5,500 steps per day for improved health related quality of life, >7,000 steps per day for better immune function, and 8,000 to 10,000 steps per day for an effect on metabolic syndrome and maintenance of weight (Scottish Government, 2019). NR: no response

Figure 2 shows the barriers and facilitators that were identified during patient interviews, grouped by COM-B component, and Figure 3 shows those identified in stakeholder interviews.

**Figure 2:**
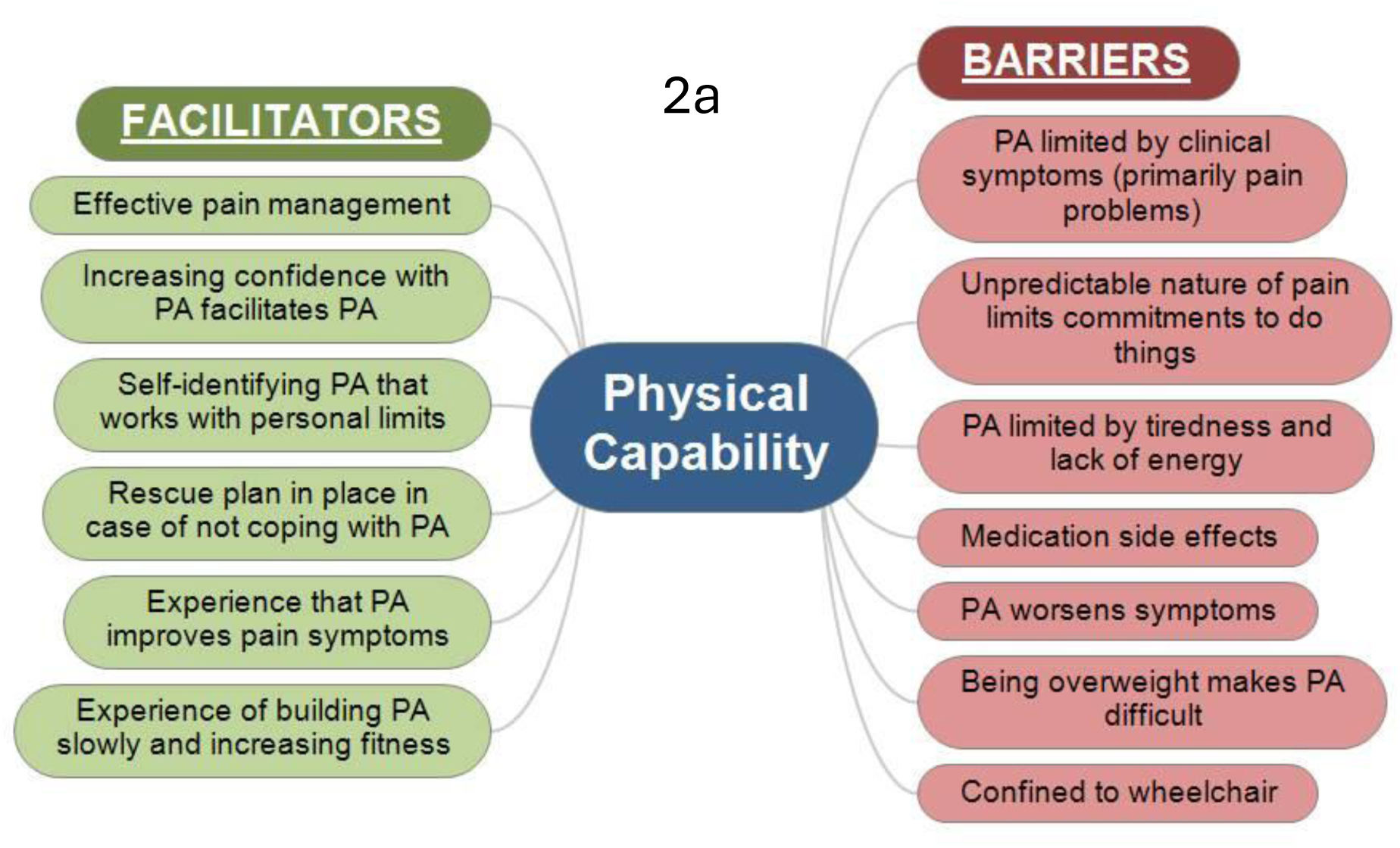
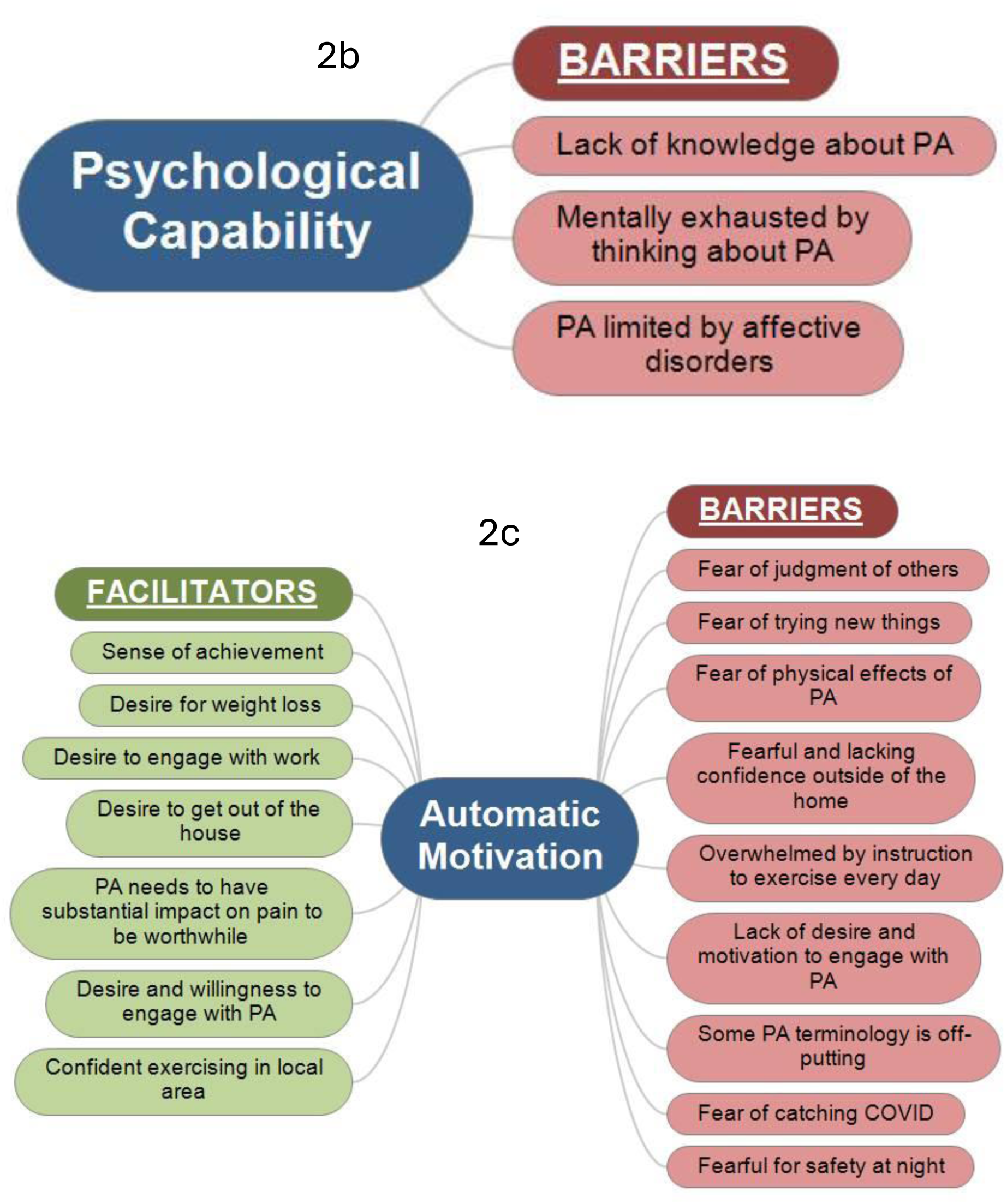
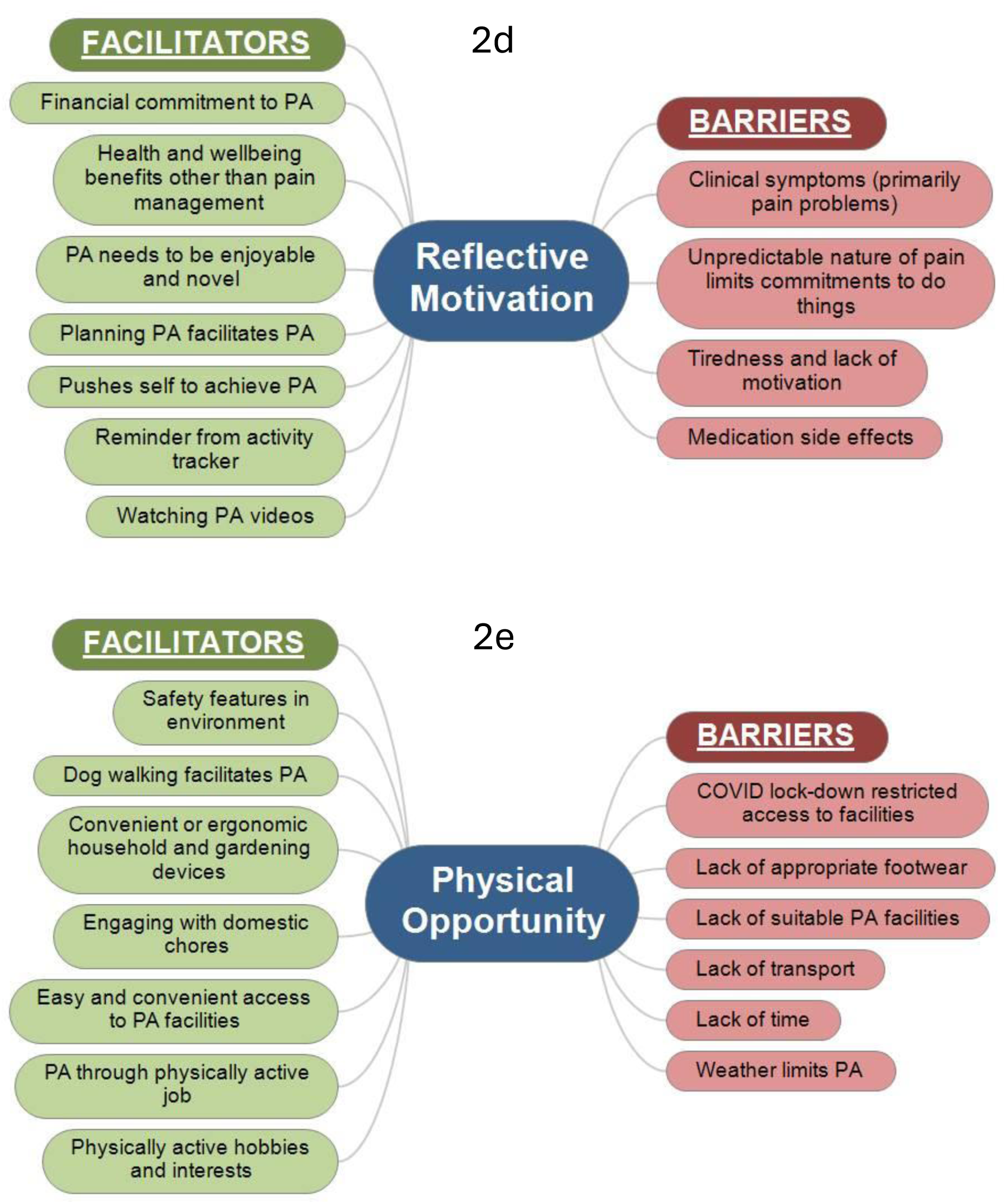
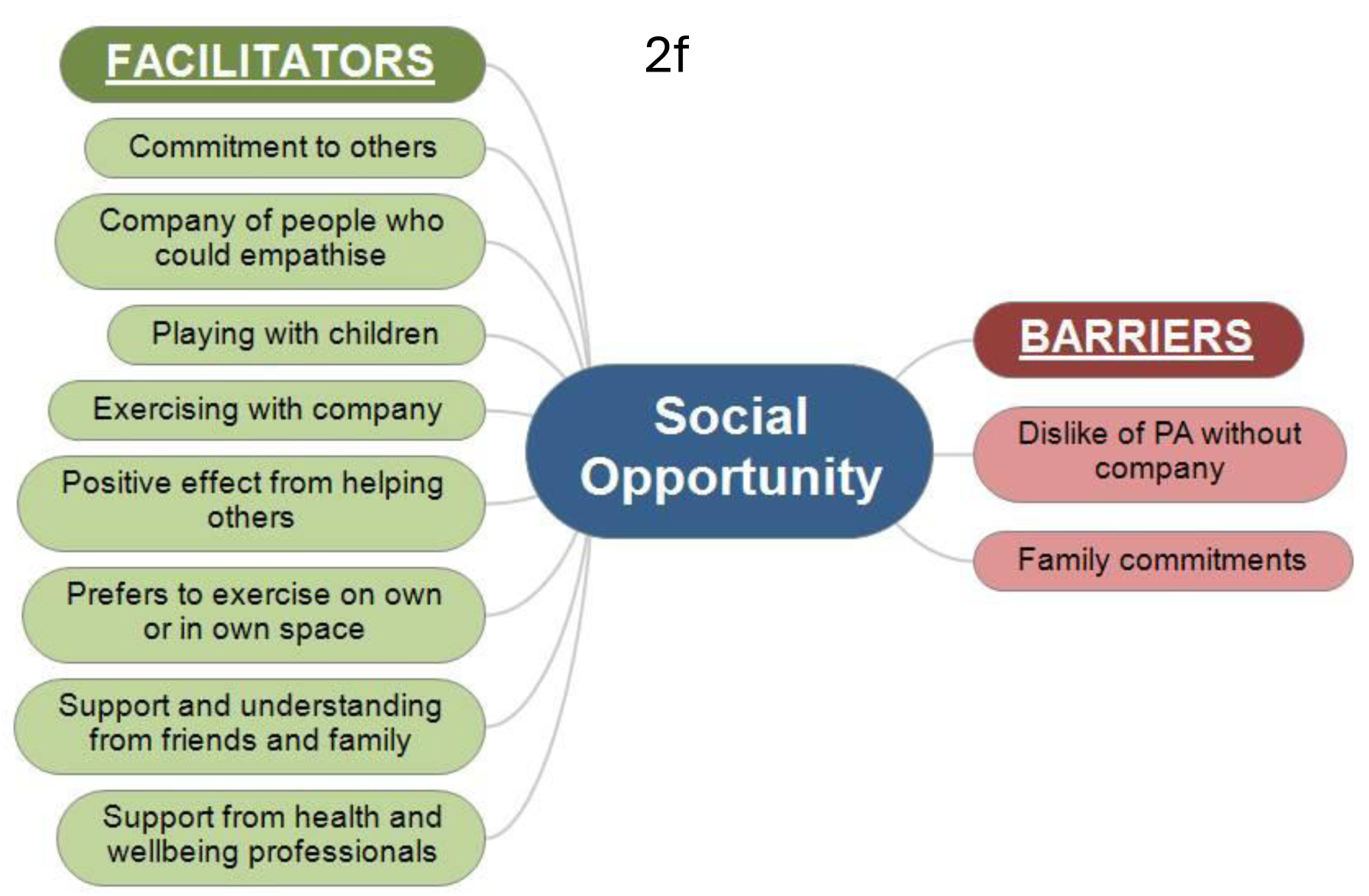
Patient-reported barriers and facilitators to engaging with physical activity in people with chronic pain

**Figure 3:**
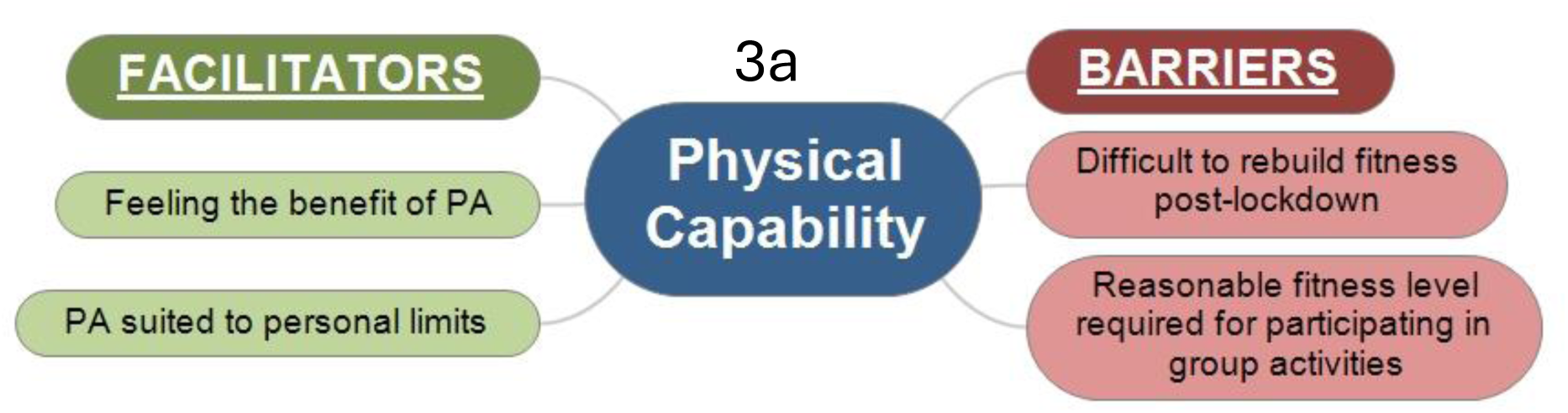
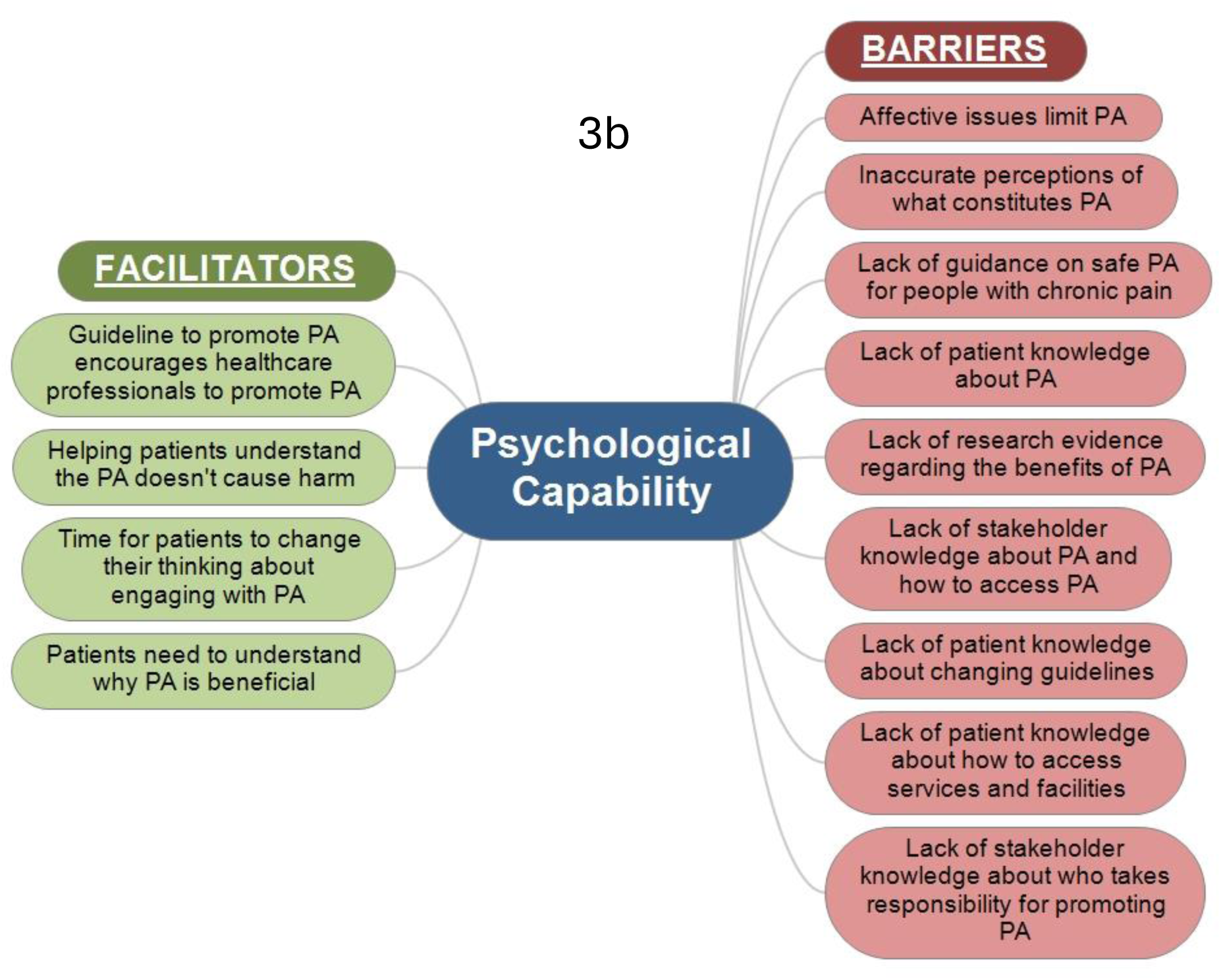
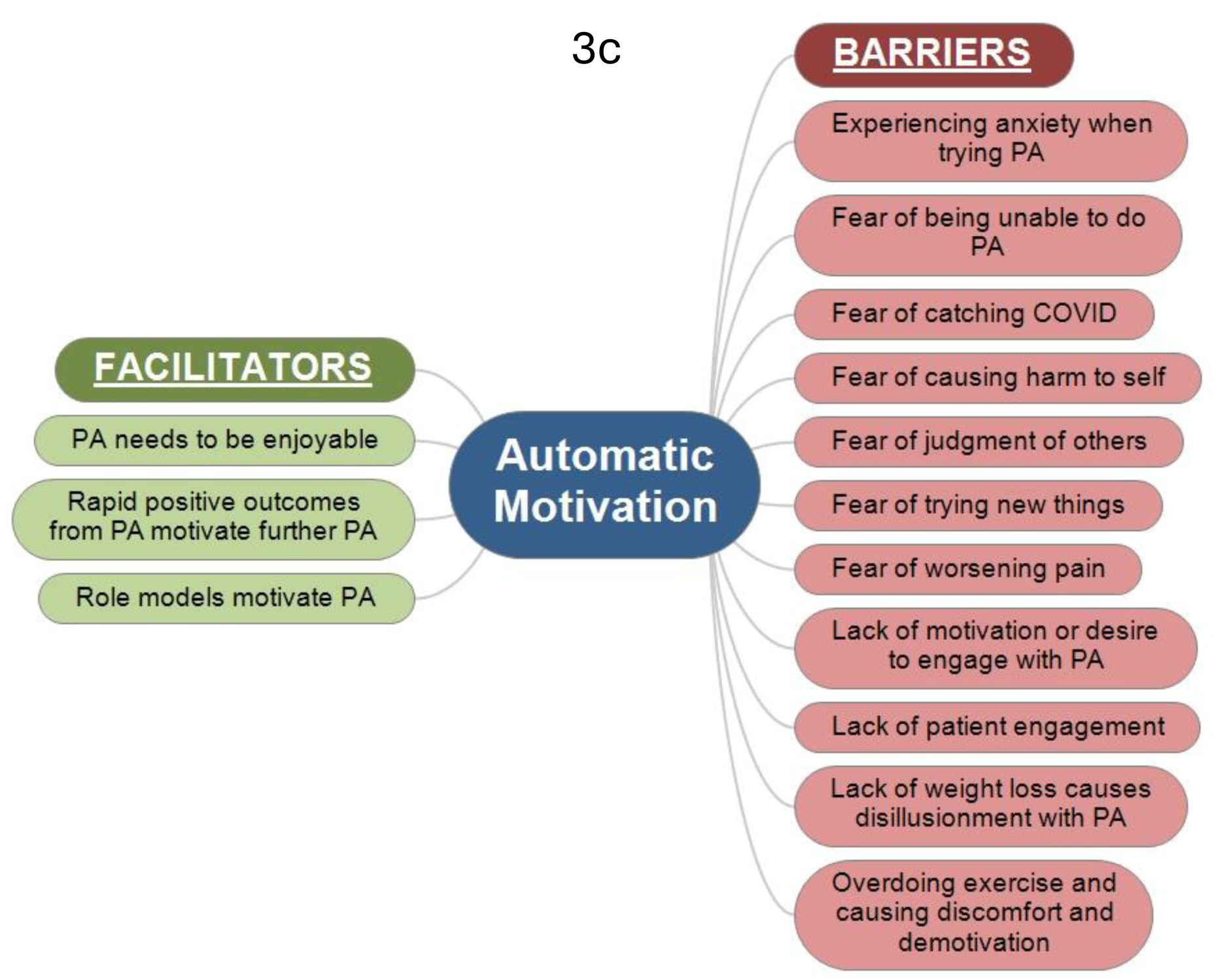
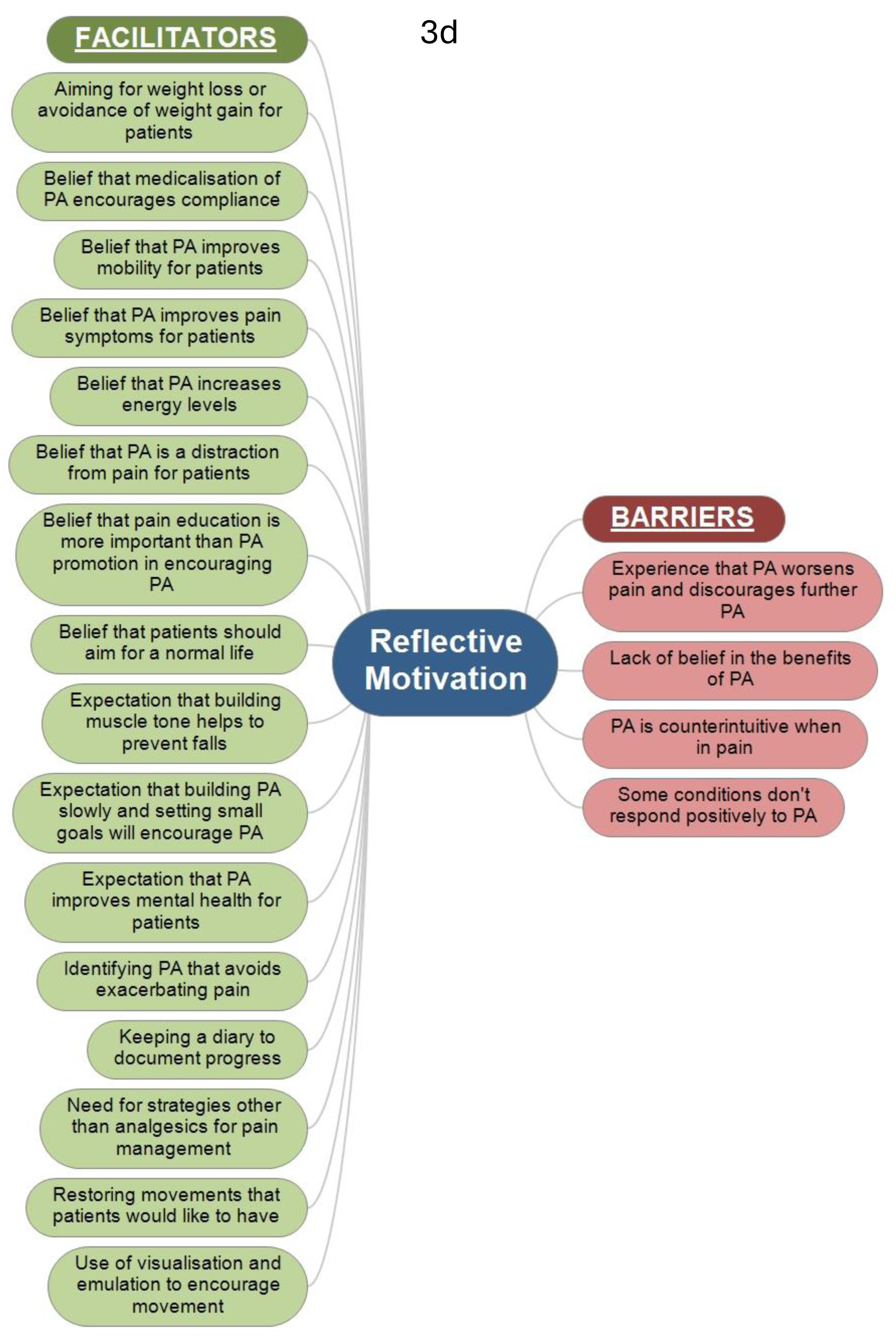
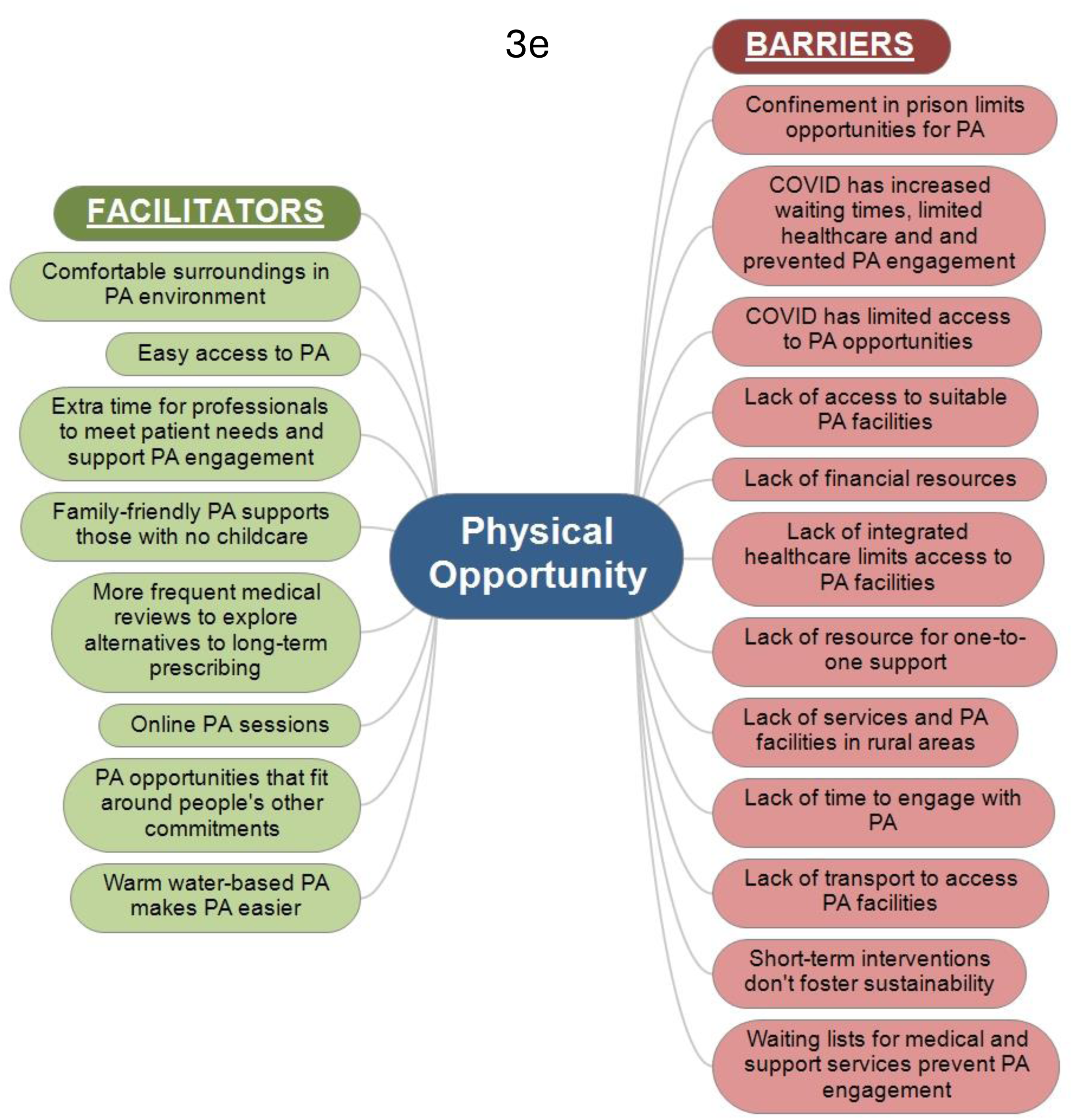
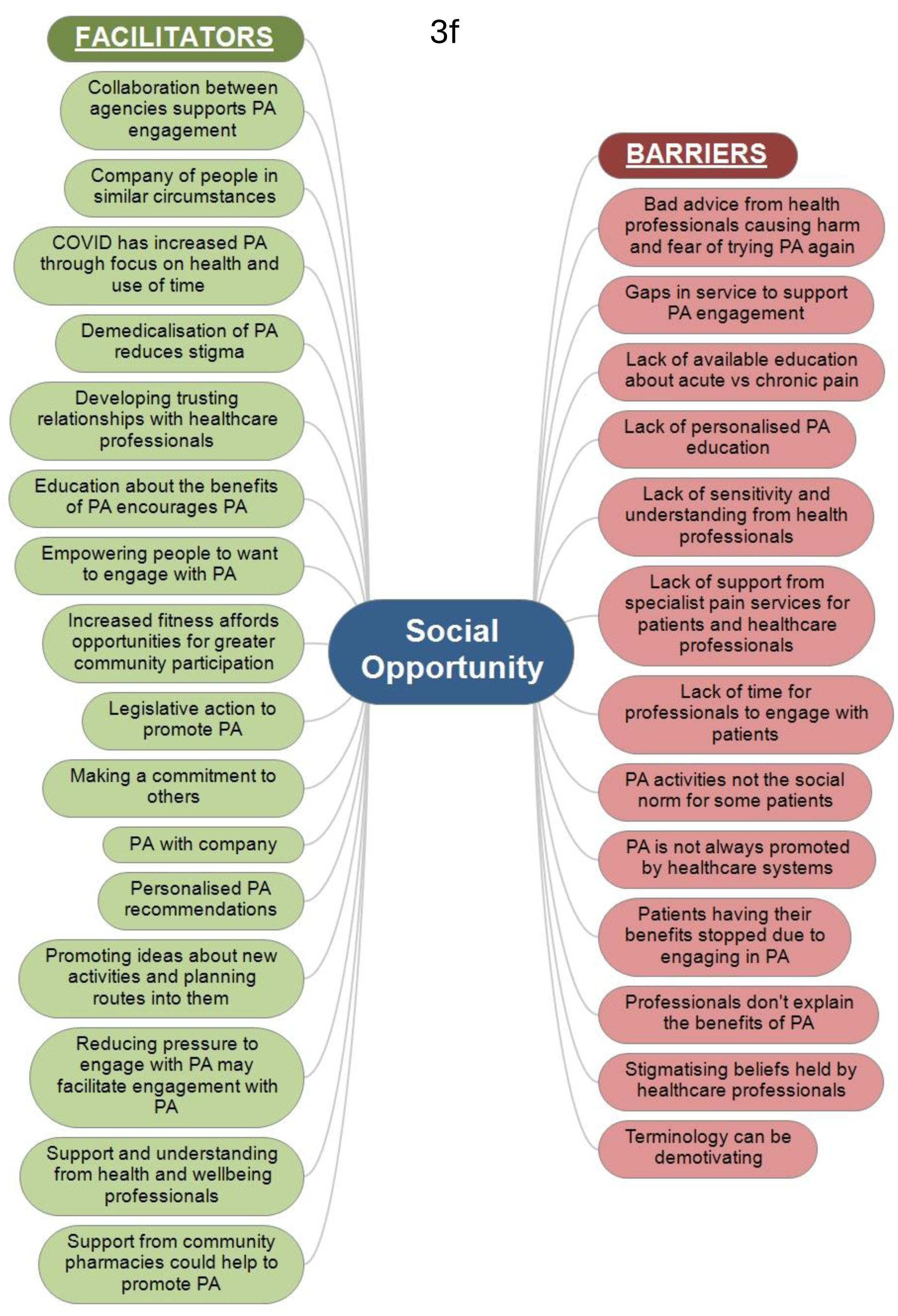
Stakeholder-perceived barriers and facilitators to engagement in physical activity by patients with chronic pain

An overview of the prominent barriers and facilitators to PA identified by people with CP and stakeholders is presented in the remainder of this section, organised by the components of the COM-B framework. Quotes from patients are coded either ‘Axx’ or ‘Fxx’, and quotes from stakeholders are coded ‘Sxx’.

### Capability – physical

Patients reported that PA was limited by physical symptoms, primarily prohibitive pain intensity, and that the pain experience worsened when performing PA:

> I used to love taking the dog out for a walk and now I just can’t do it, I can’t even walk down to the bottom of the road where the paper shop is because it’s just too painful [A12]

> … whenever I do the garden, I know when I’m doing it and I know the next day I’m going to be in horrific pain and it’s going to be a really bad day [A11]

Patients also felt that the unpredictable nature of pain prevented them from committing to participating in activities because failing to honour commitments impacted either on them or on others:

> Planning is very, very difficult, it’s made a massive inroads into both myself and my wife’s lives, it’s very difficult to plan anything with any certainty at all [A16]

Patients reported that effective pain management was an important facilitator for engaging with PA:

> I just needed something to give me that wee bit of pain relief so I could cope in the mornings and do stuff, get up, get a shower [F08]

and many felt that PA could achieve this effectively:

> I find walking helps as well. If I go out on slow walks it just helps the pain if I do that but then it is painful as I’m walking but the pain seems to ease as I’m walking [A09]

Additionally, for some, having a rescue plan in place facilitated PA engagement:

> It’s like I’ve got a wheelchair if need be and so I’d give it a go and would take that as a back up [A08]

### Capability – psychological

Both groups felt that affective symptoms limited PA:

> I’ve got lack of motivation anyway because of depression [A12]

> Mental health, that is a massive barrier [S02]

and stakeholders highlighted the dynamic relationship between pain and mental health, which further limits opportunities for PA:

> A lot of people haven’t been out of the house for so long due to their pain they almost develop a secondary social condition where they just become really sort of agoraphobic [S11]

However, the most prominent issue was lack of patient and stakeholder knowledge about the benefits of PA and how to access PA opportunities:

> I would be able to do things but with no knowledge to be able to do more exercise I can’t do it. [F17]

> I probably wouldn’t recommend anybody going to the gym and that’s probably around my lack of knowledge [S06]

Stakeholders observed that the availability of a PA guideline encouraged healthcare professionals to promote PA to patients but that patients need time to make mental shifts in how they think about PA. They also observed that patients need to understand why PA is beneficial and that PA does not cause harm in people with CP:

> [Exercise is] more beneficial when you know why you’re doing it … [and understand that] this movement isn’t going to cause me any damage, but it will take time [S02]

### Motivation – automatic

First, both groups reported fear of the physical effects of PA, largely worsening pain or causing harm, as a prominent barrier:

> Sometimes I have concerns that the exercise I’m doing, although I pace myself, might be doing damage. [A14]

> If you do something that hurts, part of the frustration is the pain at the time, and part of the worry is how bad is it going to be in 24 hours’ time or 40 hours’ time? [S12]

Secondly, participants in both groups reported a lack of patient desire and motivation to engage with PA:

> The actual motivation to go and do it, even though it’s only going to maybe take a couple of minutes, the actual motivation to go out and do it, no. [A12]

> I think it takes a lot more motivation for someone with chronic pain to exercise than someone that doesn’t have chronic pain. [S12]

Thirdly, fear of the judgement of others was seen as a substantial barrier by both groups, with a focus on physical signs of health conditions (such as a hernia or skin condition) and lack of capacity for PA:

> There is exercise class but they do a lot of things I couldn’t do. Somebody said to me to sit in a chair and do some of them but without anybody else sitting in a chair and doing exercises I would feel out of place or looked at or something. [A06]

Three prominent facilitators were identified by patients: desire and willingness to engage with PA; desire for weight loss; and desire to get out of the house. Stakeholders failed to recognise desire and willingness in patients; however, it was widely expressed by patients:

> So if there’s something that is clinically proven and something that’s worth trying, I would definitely give it a try, whatever form of exercise that may be. So I’m open to that. Absolutely. [F06]

### Motivation – reflective

Patients reported one prominent barrier: lack of trust in PA guidance:

> There’s been some days I’ve thought whoever thought that up [PA improves pain] must have been off their head because it’s not true [A05]

and stakeholders reported one prominent barrier: the experience that PA worsens patients’ pain and discourages further PA:

> … psychologists often feel that if we’re going to encourage [PA] that are we at risk of causing further worsening chronic pain. [S10]

> … they try and do some exercise and it will be sore so that puts them off persevering with that. [S08]

The most prominent facilitator reported by patients was benefits to physical and mental health, and the stakeholders focused specifically on the benefits to mental wellbeing:

> I know that the exercise is necessary to avoid diabetes and muscle issues and all of the other stuff. [F02]

> There is obviously the strength, the fitness but also the feel-good hormones and chemicals that are produced in the brain that help us feel better [A07]

> … the [physical health] benefits that you’re going to get are almost secondary to the fact that you’re able to get out there for your mental health. [S01]

An additional prominent factor reported by patients was that PA needs to be enjoyable and novel:

> And what works for me because I don’t want to push myself, I want to actually enjoy the exercise to get myself back into it. [F15]

Additionally, stakeholders believed that building PA slowly and setting small goals would encourage PA:

> It working with the patient to help them find a level that is a baseline and work their way up that’s going to be beneficial and not knock them back even further than they were before. [S10]

### Opportunity – physical

The most prominent barrier reported in both groups was restricted access to PA opportunities due to COVID-19 lockdowns, which is likely to have been a transient situation, but they also reported a lack of PA facilities more generally. Patients also reported the impact of bad weather and a lack of time, and stakeholders highlighted lack of financial resources for patients:

> Weather is a demotivating or a motivating factor. [A25]

> Yes, time because it’s obviously half an hour there, half an hour back which is a really big chunk of time that I could be trying to get through tasks on my to do list. [A07]

> … if you Google it and see there’s nothing there then what’s the point in saying you know why don’t you go to so and so. They can only be done during the day and I work so.

> The cost, there’s a lot of things out there with exercise that’s quite expensive. [S03]

Many of the facilitators identified by patients related to chance opportunities, such as having a physically-active job, engaging with domestic chores or dog-walking activities. They also reported that having easy and convenient access to PA facilities encouraged PA:

> So I thought if I go to a gym, there’s one that’s not far from me, if I’m tired, I could get a bus there, you know what I mean? I’ve got some way to get there and get home. [F08]

Stakeholders suggested a number of additional potential facilitators that were not reported by patients, including making use of online PA classes and the provision of family-friendly PA to support those with no childcare:

> One of the things that we’ve noticed now we’ve got some family classes now where if somebody’s just had a kid or they’ve got a couple of children … they [children] could come in and take part in the classes or do something at the same time. [S01]

### Opportunity – social

Patients reported two barriers: dislike of PA without company (lack of enjoyment, lack of confidence and lack of motivation to persevere); and family commitments limiting opportunities for PA.

> It is something that I would be interested in, but as I say I’m just not confident to go myself. [A12]

> Sometimes when the kids are being really difficult and just want to fight with each other all the time and I feel like I can’t just go out and do the garden … I need to be in the same room as them to make sure they’re not at each others’ throats [A07]

Stakeholders identified numerous additional barriers that related to both issues for patients (PA activities not the social norm for some patients; patients having their benefits stopped due to engaging in PA; and the demotivating effect of some terminology) and concerns around the limitations of service provision (gaps in provision and unhelpful attitudes or stigmatising beliefs held by professionals):

> [Patients] struggle a lot to be validated and believed … so if you go and say something so crass as go exercise, all their fears have been confirmed in that one statement, you have no idea what is going on with me because if you did you would never have said that in the first place. [S11]

> I think in health care in particular, it’s a lot of judgement. [S05]

The most prominent facilitators identified by patients were support from friends and family and support from health professionals, with stakeholders agreeing with the latter. The other prominent facilitators, identified by both groups, concerned PA with company and, especially, PA in the company of people who could empathise with their difficulties. Stakeholders proposed additional potential facilitators that concerned both service development (enhanced collaboration between agencies and community pharmacy involvement in promoting PA engagement) and the direct patient experience (largely characterised by the provision of education and support to encourage sustainable PA and the way in which professionals engage with patients):

> That’s sometimes where building a relationship with the patient helps, though, because once they trust you and they kind of know that you’re looking out for them to push it a bit more and say, what have you thought about x, y and z? … You can get the confidence to have those kind of conversations just because you know the person better. [S13]

## Discussion

### Summary

Many of the barriers and facilitators that were identified by both groups and by the patient group only echoed previously reported findings, providing further evidence of their importance and validity. However, the patient group raised additional barriers not already addressed in the existing literature that highlight a more complex range of barriers relating specifically to physical capability and automatic motivation. Based on their clinical experience, stakeholders raised an inventory of barriers that had not been raised by participants in the patient group, spanning reflective motivation, physical opportunity and social opportunity. Regarding social opportunity, stakeholders identified additional barriers that related to both issues for patients and concerns around the limitations of service provision. The stakeholders also observed a range of facilitators, within the domains of psychological capability, reflective motivation, physical opportunity and social opportunity, that had already proven beneficial for some of their patients. Regarding social opportunity, they proposed facilitators that concerned both service development and the direct patient experience.

### Strengths and limitations

The present study examined both barriers to and facilitators of PA in a sizeable sample of people living with clinically significant CP, with additional contributions from the perspective of stakeholders, using a robust behavioural change model to understand the data with a view to identifying interventions that could increase PA engagement and activity levels. Researcher preconceptions can result in the potential for bias; however, to mitigate this, an interdisciplinary team was established, and team members were intensively involved in developing the analytical framework and interpreting the findings both inductively and deductively.

### Comparison with existing literature

There is a considerable literature examining the facilitators and barriers to PA in people with chronic, painful conditions; however, there is only a limited literature that frames these facilitators and barriers within a robust framework that could be used to affect behavioural change. A recent rapid umbrella review (Webb *et al*., 2022) included 12 systematic reviews that framed barriers and facilitators within the COM-B model for people with musculoskeletal conditions (largely various types of arthritis); however, half used quantitative methods, which may have restricted the richness of information elicited from participants. The barriers and facilitators reported in this rapid review were similar to many of the prominent ones identified in the present study providing further evidence of their key significance, including: prohibitive pain intensity and activity-induced exacerbation of pain (physical capability); lack of knowledge about the health benefits and analgesic effects of PA (psychological capability); fear of causing further harm and the unpredictability of pain symptoms preventing habit formation (automatic motivation); negative beliefs and experiences of PA on pain symptoms (reflective motivation); inaccessible facilities/activities and deterred by lack of time or bad weather (physical opportunity); and lack of company when performing PA (social opportunity).

However, the present study identified a greater range of patient-reported barriers than have been identified in previous studies, including, for example: weight problems making PA difficult; sedating side effects of medication; and fear of the judgement of others. Identifying an exhaustive range of barriers is important when considering developing inclusive interventions that have the capacity to anticipate and address the needs of all patients, reduce inequalities and support behavioural change. The inclusion of stakeholder data in the present study elucidated further, describing barriers that had not been identified by patient participants. For example, whilst it is often presupposed that healthcare providers will assume responsibility for PA promotion, stakeholders also face barriers related to psychological capability (e.g. lack of knowledge about PA and lack of knowledge around who should assume which responsibilities), which are barriers that were also identified in a recent narrative review (Leese *et al*., 2023). Stakeholders also reported barriers relating to physical opportunity that were faced by patients due to limitations in healthcare systems (e.g. excessive waiting times reducing access to physiotherapy and other healthcare services), and they identified several barriers relating to social opportunity concerning interactions with patients (e.g. lack of their own time to engage with patients and lack of sensitivity and understanding from professionals); limitations in healthcare systems (gaps in service provision and lack of support from specialist services); and PA activities not being the social norm for some patients.

### Implications for research and practice

The identification of important factors in all three components of the COM-B model suggests that behavioural change interventions must address multiple aspects. Whilst addressing one component of COM-B has the potential to impact positively on the others, it is likely that maximum effect would be achieved by increasing all three: capability; opportunity; and motivation. The findings of the present study suggest that consideration of these three components, and precisely identifying those relevant to individuals, could assist healthcare practitioners in developing a more tailored approach to assessing the barriers encountered by individuals and promoting PA engagement and increased activity levels in people with CP.

## Conclusion

The barriers and facilitators relevant to PA in people with CP are numerous and complex. The COM-B framework provides a mechanism for organising and understanding these barriers and facilitators in a way could that inform intervention design. Future research should develop and evaluate such interventions with the aim of promoting PA engagement and increasing activity levels in people with CP.

## Data Availability

All data produced in the present study are available upon reasonable request to the authors.

## Funding

The study was funded by the Chief Scientist Office (ref: CSO HIPS/19/26).

## Ethical approval

The study was approved by the London - Chelsea Research Ethics Committee (reference number: 19/LO/2012).

## Competing interests

The authors have no competing interests to disclose.

## Acknowledgements

We would like to acknowledge the Chief Scientist Office for having funded this study (ref: CSO HIPS/19/26). We would also like to acknowledge the contribution made by the participants to this study and the Tayside Clinical Trials Unit (TCTU), University of Dundee for its role in study management.

## Notes

### Competing Interest Statement

The authors have declared no competing interest.

